# Improved diagnosis of inflammatory bowel disease and prediction and monitoring of response to anti-TNF alpha treatment based on measurement of signal transduction pathway activity

**DOI:** 10.1101/2022.05.16.22275125

**Authors:** Wilbert Bouwman, Wim Verhaegh, Anja van de Stolpe

## Abstract

**Objective:** Ulcerative colitis (UC) and Crohn’s disease (CD) are two subtypes of chronic inflammatory bowel disease (IBD). Differential diagnosis remains a challenge. Anti-TNFα treatment is an important treatment for IBD, yet resistance frequently occurs and cannot be predicted. Consequently, many patients receive ineffective therapy with potentially adverse effects. Novel assays are needed to improve diagnosis, and predict and monitor response to anti-TNF-α compounds.

**Design:** Signal transduction pathway (STP) technology was used to quantify activity of STPs (androgen and estrogen receptor, PI3K, MAPK, TGFβ, Notch, Hedgehog, Wnt, NFκB, JAK-STAT1/2, and JAK-STAT3 pathways) in colon mucosa samples of CD and UC patients, based on transcriptome analysis. Previously described STP assay technology is based on computational inference of STP activity from mRNA levels of target genes of the STP transcription factor.

**Results:** Results show that NFκB, JAK-STAT3, Wnt, MAPK, and androgen receptor pathways were abnormally active in CD and UC. Colon and ileum-localized CD differed with respect to STP activity, the JAK-STAT1/2 pathway being abnormally active in ileal CD. High activity of NFκB, JAK-STAT3, and TGFβ pathways was associated with resistance to anti-TNFα treatment in UC and colon-located CD, but not in ileal CD. STP activity decreased with successful treatment.

**Conclusion:** We believe that measuring mucosal STP activity provides clinically relevant information to improve differential diagnosis of IBD and prediction of resistance to anti-TNFα treatment in patients with colon-localized IBD, and provides new targets for treatment and overcoming anti-TNFα resistance.

## Introduction

Inflammatory bowel disease (IBD) is an inflammatory chronic intestinal disorder. The two major types include ulcerative colitis (UC) and Crohn’s disease (CD). Crohn’s disease affects the small intestine and large intestine, as well as the mouth, esophagus, stomach and anus, whereas ulcerative colitis primarily affects colon and rectum. Differential diagnosis with respect to CD and UC remains a challenge, and in 15% of cases, definitive discrimination is not possible. Correct diagnosis is important to decide on the most suitable therapy including systemic drug treatment. Systemic therapy is directed at inhibition of inflammation and the auto-immune process, healing of the intestinal wall, and correction of the microbiome ^1^. Multiple types of drugs can be used, e.g. aminosalicylic acids, corticosteroids, and tumor necrosis factor alpha (TNFα) blockers, but around one third of patients will be primary resistant to therapy and three quarters of responders develop secondary resistance within one year of treatment ^2^,^3^,^4^,^5^. As a consequence, many patients receive ineffective therapy with potentially severe adverse effects, including serious infections. Unfortunately, biomarker assays to predict response to anti-TNF-α drugs, to measure the extent of the response, and to identify emerging drug resistance are lacking, precluding a personalized treatment approach ^6^. Novel assays to improve diagnosis, and to predict and monitor response to anti-TNF-α compounds are needed. New drug targets are also needed for those patients who are resistant to conventional treatment, including anti-TNFα therapy.

We show that our signal transduction pathway (STP) technology can be used to develop these clinically needed assays, and to identify new drug targets in patients with CD and UC.

## Methods

### Measurement of activity of signal transduction pathways on Affymetrix microarray data from cell samples

Development and validation of computational models to quantitatively measure activity of the androgen receptor (AR), MAPK, NFκB, WNT, TGFβ, JAK-STAT1/2, and JAK-STAT3 pathways have been described before, and can in principle be used to analyze pathway activity on any cell type, based on analysis of transcriptome data ^7^,^8^,^9^,^10^,^11^,^12^,^7^,^13^,^14^. For the current study, pathway activity scores (PAS) were measured on publicly available Affymetrix expression microarray data from preclinical and clinical IBD studies.

PAS were calculated from Affymetrix expression microarray data obtained from the GEO database^15^. For each signaling pathway, PAS represent the log2 of the odds of the pathway-associated transcription factor being active vs. inactive as calculated by a probabilistic computational model, as described before ^8^,^12^,^9^.

### Microarray Data Quality Control

Quality control (QC) was performed on Affymetrix data of each individual sample, using standard methods as described before ^9^. In summary, QC parameters include: the average value of all probe intensities, presence of negative or extremely high (> 16-bit) intensity values, poly-A RNA (sample preparation spike-ins) and labelled cRNA (hybridization spike ins) controls, *GAPDH*, and *ACTB* 3’/5’ ratio, center of intensity and values of positive and negative border controls determined by affyQCReport package in R, and an RNA degradation value determined by the AffyRNAdeg function from the Affymetrix package in R ^16, 17^. Sample data that failed QC were removed prior to data analysis.

### Brief description of the analyzed preclinical and clinical studies

#### GSE16879 ^18^

Mucosal biopsies had been obtained at endoscopy from actively inflamed mucosa from IBD patients (24 ulcerative colitis (UC), 19 Crohn’s colitis (CDc) and 18 Crohn’s ileitis (CDi)), refractory to corticosteroids and/or immunosuppression, before and 4-6 weeks after intravenous infliximab (5 mg/kg) infusion, and from normal mucosa of 12 control patients. Biopsies had been taken at sites of active inflammation; in case of healing the repeat biopsy had been obtained in the area where lesions had been present prior to anti-TNFα therapy. Clinical response had been assessed 4-6 weeks after starting treatment. Of note, response assessment criteria used for CDc had been adapted to define (partial) response in CDi.

#### GSE23597 ^19^

48 UC patients with moderate-to-severe UC, refractory to corticosteroids, had been treated with 5 or 10 mg/kg infliximab or placebo at weeks 0, 2, 6 and every 8 weeks thereafter. Colon biopsy samples had been collected 15 to 20 cm from the anal verge prior to treatment and during treatment at weeks 8 and 30, that is, after induction and during maintenance therapy, respectively.

#### GSE52746 ^20^

Colon mucosa biopsy samples had been taken from CD patients and 17 non-inflammatory controls. CD patients included 10 patients with active CD without anti-TNF therapy, 5 with active CD under anti-TNF therapy (non-responders) and 7 patients with inactive CD under anti-TNF therapy (responders). In patients with active CD, biopsies had been taken from the affected areas; in patients with inactive disease from the healed region. Patients had been either treated with adalimumab or infliximab. Adalimumab had been given subcutaneously (160/80 mg) at weeks 0 and 2 for induction, and (40 mg) every other week for maintenance treatment. Infliximab had been given intravenously (5 mg/kg) at weeks 0, 2, and 6 for induction, and every 8 weeks for maintenance treatment. For six patients paired biopsy data were available before and after 12 weeks of anti-TNFα treatment (5 responder patients and one non-responder).

### Privacy

All analyzed data were obtained in agreement with GDPR regulations, as implemented within Philips.

### Interpretation of signal transduction pathway activity score (PAS)

An important and unique advantage of the STP technology is that in principle analysis can be performed on each cell type. As described before ^21^, one should bear in mind a few considerations when interpreting log2 odds PAS:

1. On the same sample, log2 odds PAS cannot be compared between different signaling pathways, since each of the signaling pathways has its own range in log2 odds activity scores ^21^.
2. The log2 odds range for pathway activity (minimum to maximum activity) may vary depending on cell type. Once the range has been defined using samples with known pathway activity, on each new sample the absolute value can be directly interpreted against that reference. If the range has not been defined, only differences in log2 odds activity score between samples can be interpreted.
3. PAS are highly quantitative, and even small differences in log2 odds PAS can be reproducible and meaningful.
4. A negative log2 odds ratio does not necessarily mean that the pathway is inactive.

### Statistics

Boxplots, individual sample plots and hierarchical clustering plots have been made using the Python data visualization library function seaborn; additional statistical annotations have been created using Python package statannot. Box-and-whisker plots show median values, interquartile range (IQR) between 25^th^ and 75^th^ percentile, extreme values, and outlier samples (diamond shape). Individual samples are presented as small circles (outlier samples are hence plotted twice, as diamond and as circle). Two-sided t-test (low sample numbers) or Mann-Whitney-Wilcoxon testing was used to compare PAS across groups. P-values are indicated in the figures, *with p<0*.*01 considered as significant*. Mean log2 odds, mean delta between groups, P-values, u-stats and area under the curve (AUC) values have been summarized in Supplementary Table 1.

## Results

STP activity scores from colon mucosa biopsy samples from clinical studies on CD and UC are shown in Figures 1-4 and Supplementary Figures S1 and 2). In the description of results, unless otherwise specified, mentioned differences in PAS are significant, meaning p<0.01.

**Figure 1.**
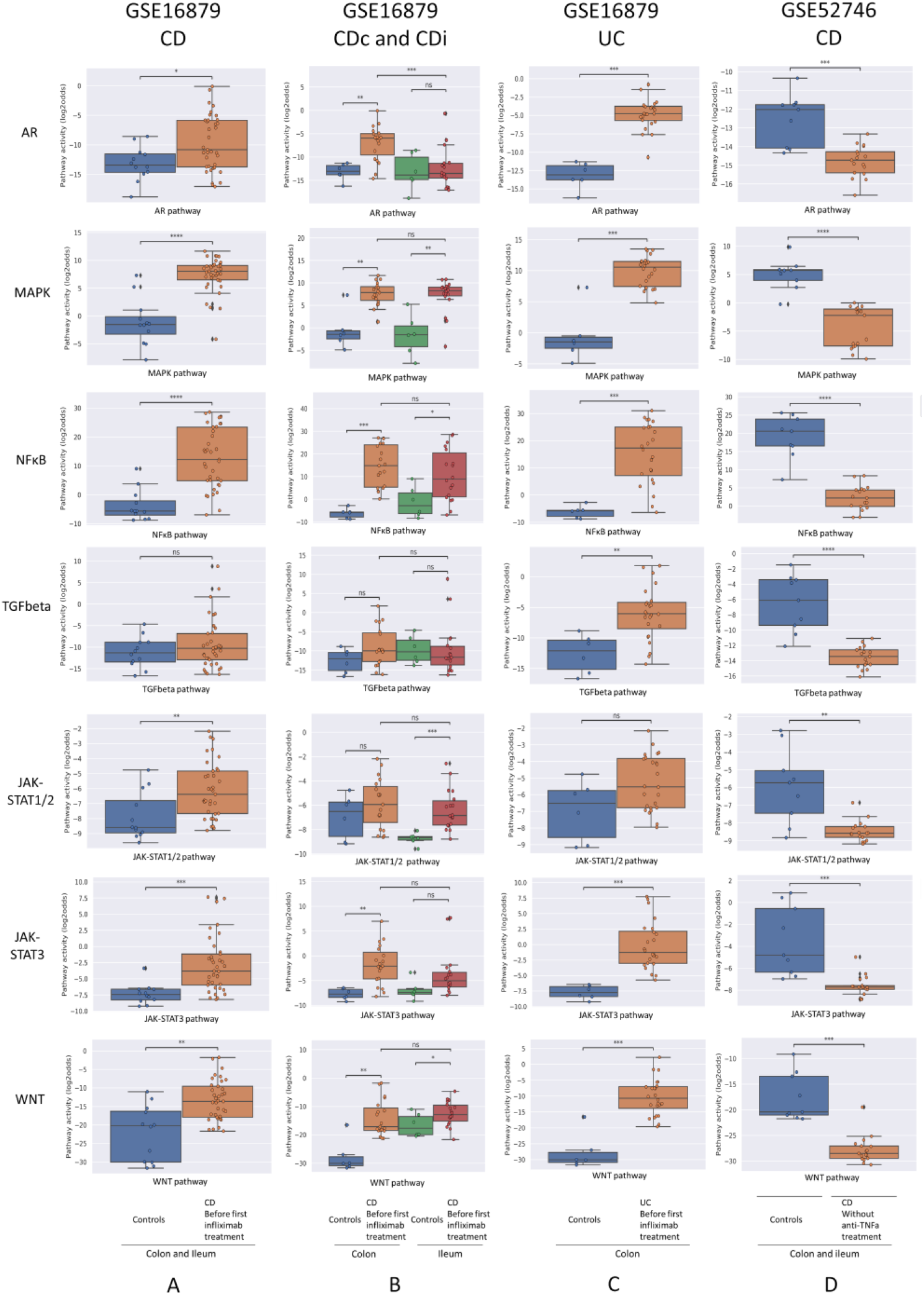
Diagnosis of IBD. Datasets indicated on top of figure (A-D). STP activity analysis of intestinal mucosa samples for diagnosis of IBD. Mucosa samples from healthy control individuals and pre-treatment mucosa samples from IBD patients. For details, see Methods. Signal transduction pathway (STP) activity scores (PAS) are shown for the androgen receptor (AR), MAPK, NFκB, TGFβ (TGFbeta), JAK-STAT1/2, JAK-STAT3, and Wnt STPs. PAS on Y-axis on a log2 odds scale. Two sided Mann-Whitney-Wilcoxon statistical tests were performed; p-values are indicated in the figures as *p < 0.05, ***p < 0.001, ****p < 0.0001, ns: not significant.

### Diagnosis of CD and UC

To investigate which STPs are abnormally active in colon mucosa of IBD patients, and whether measuring STP activity can improve diagnostic accuracy, and can be used to facilitate differential diagnosis of CD and UC, STP activity was calculated for mucosa biopsy samples of IBD patients with different clinical manifestations. For CD, colon biopsy sample data were available from patients with specific involvement of either colon (CDc) or ileum (CDi) (dataset GSE16879), and of patients with disease present in colon, but not necessarily limited to colon (dataset GSE52746).

In untreated CD (dataset GSE16879, CDc and CDi combined), MAPK, NFκB, JAK-STAT1/2, JAK-STAT3, and Wnt PAS were increased compared to healthy controls (Figure 1A, left column, and Figure S1). There were also clear differences in pathway activity between CDc and CDi (Figure 1B, Supplementary Table 1 and see Table 1 for summary): compared to healthy mucosa, AR, NFκB, JAK-STAT3, and Wnt PAS were increased in CDc, while JAK-STAT1/2 PAS was specifically increased in CDi. This suggests different mechanisms of disease for colon- and ileum-localized CD. Analysis of the second clinical CD study (clinical disease present in, but not necessarily limited to, colon; dataset GSE52746) showed an abnormal pathway activity profile similar to the combined profile of CDc and CDi, complemented by increased TGFβ PAS (Figure 1D; Supplementary Table 1 and Table 1 for summary).

**Table 1.**
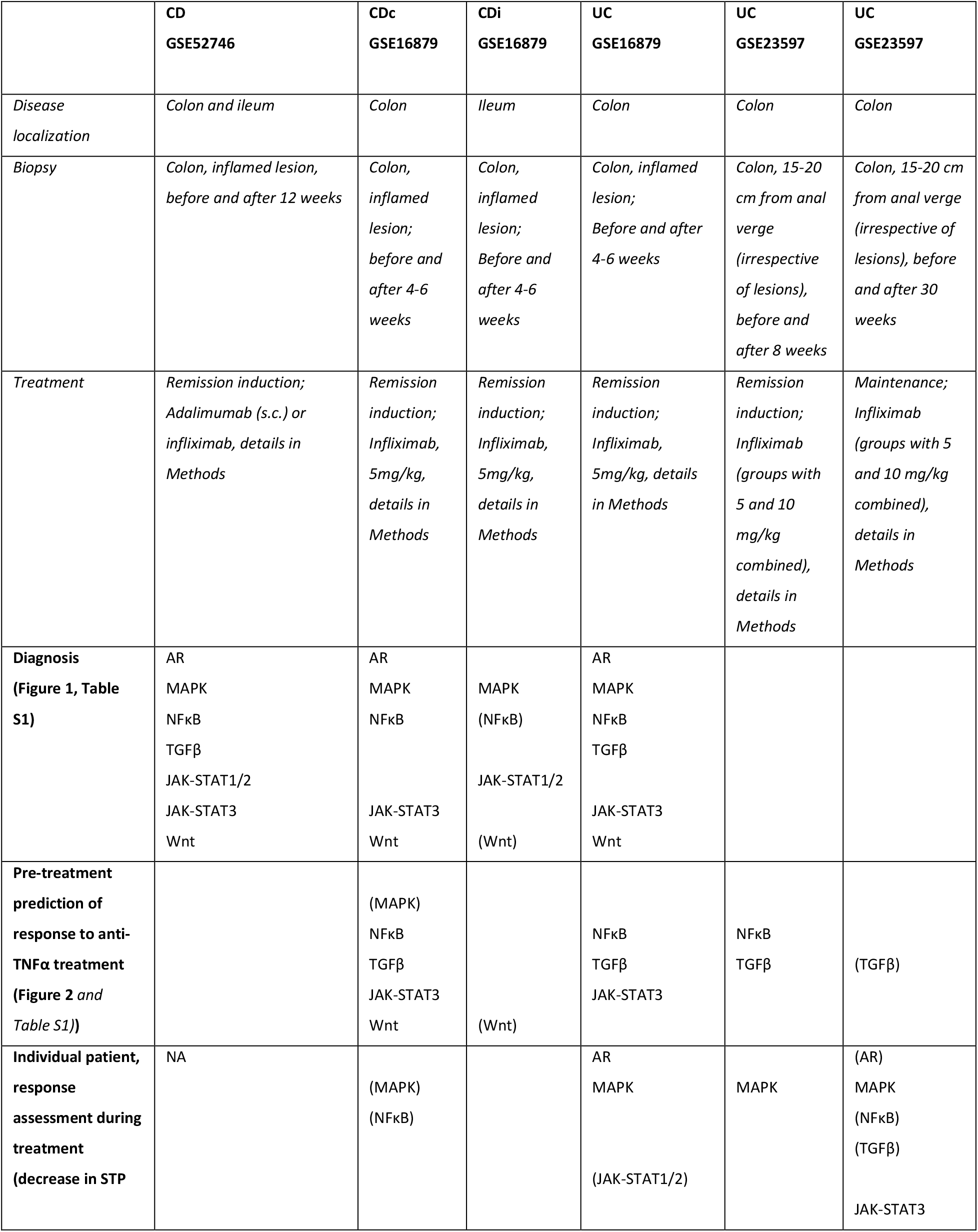
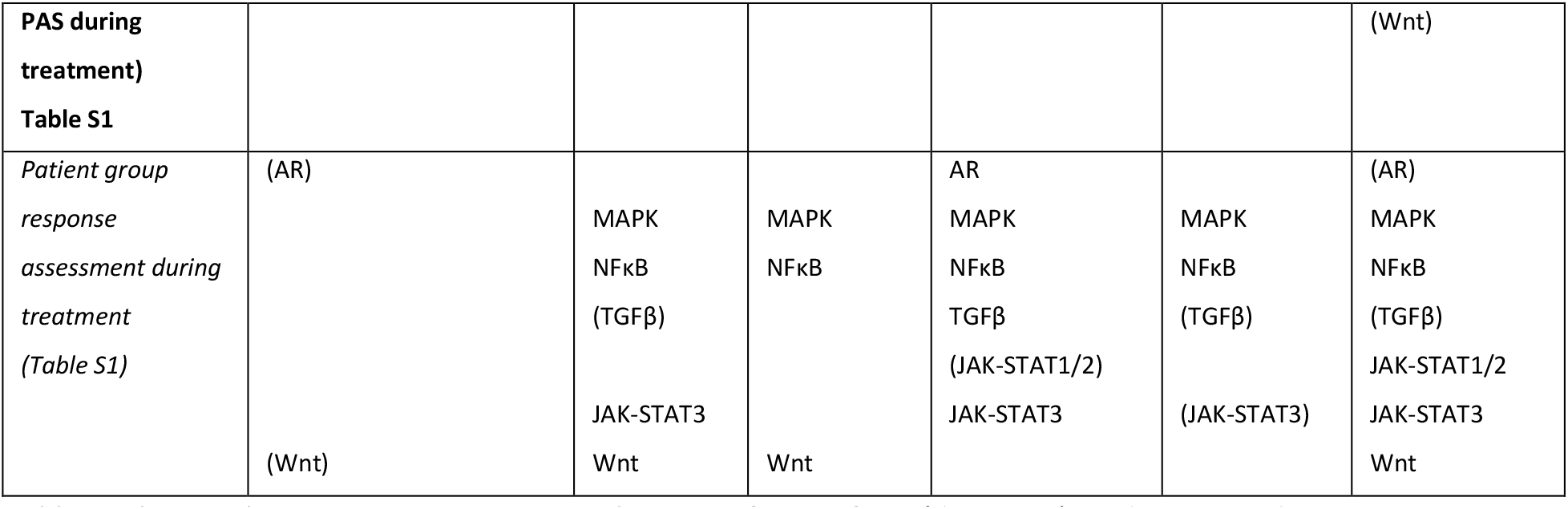
Abnormal STP activity in patients with various forms of IBD (diagnosis), and in responder versus non-responder patients, pre-treatment and during treatment. Diagnosis: comparison between IBD and healthy control STP activity; indicated STPs have higher PAS in IBD group. Pre-treatment prediction of response to anti-TNFα treatment: comparison between pre-treatment STP activity of responder versus non-responder groups; indicated STPs have higher PAS in responder patient group. Individual patient response assessment during treatment: comparison between delta (for individual patients) in STP activity before -during treatment of responder versus delta in non-responder groups; indicated STPs have higher delta PAS in responder patients. Patient group response assessment during treatment: comparison patient responder with non-responder groups during treatment; indicated STPs have higher PAS in responder group during treatment. Indicated STPs in the table: STPs with significantly higher PAS (p<0.01); between brackets: STPs with marginal significance (p<0.05). See Table S1 for absolute p-values. For specifics of patient inclusion, dosing and sample type, see Methods for description of datasets. NA: Not applicable.

For untreated UC patients (dataset GSE16879) the same STP PAS were increased as found in patients with CDc, with addition of the TGFβ pathway (Figure 1C; Table 1).

Summarizing these results (Supplementary Table 1 and Table 1 for summary), activity of the MAPK pathway was reproducibly increased in *colon* mucosa samples of patients with all investigated forms of IBD (CD, CDc, CDi, UC), indicative for a role for this pathway in the core mechanism of disease of IBD. Furthermore, IBD restricted to the ileum (CDi) was specifically associated with increased JAK-STAT1/2 pathway activity, while colon location was associated with increased AR pathway activity. Differences in STP activity between CD, CDc, CDi, and UC provide evidence for the presence of multiple IBD subtypes with variant mechanisms of disease.

### Prediction of response to anti-TNFα treatment using STP analysis of pre-treatment mucosa biopsy

To investigate whether it is possible to predict successful remission-induction by anti-TNFα treatment in IBD patients based on pre-treatment mucosa biopsy analysis, pre-treatment data from CD, CDc and CDi subtype (GSE16879; GSE527460) and UC (GSE16879, GSE23597) patients were analyzed.

In both CDc and UC patients (GSE16879), NFκB, TGFβ, and JAK-STAT3 PAS were higher in non-responder compared to responder patients (Figure 2; supplementary Table 1 and summary Table 1). CDi patients are known to generally not respond to anti-TNFα therapy, and response criteria in this group were reported to have been adapted to allow partial responders to be included in the responder group. However, no STPs were identified of which PAS predicted partial response in CDi patients. In the CD patients with colonic/ileocolonic disease from GSE52746, patients had been treated with two types of TNFα inhibitors (subcutaneous adalimumab of intravenous infliximab) and matched samples before and after treatment were only available for six adalimumab-treated patients, of which 5 responder and one non-responder patient; thus statistical analysis was not possible (Figure S2).

**Figure 2.**
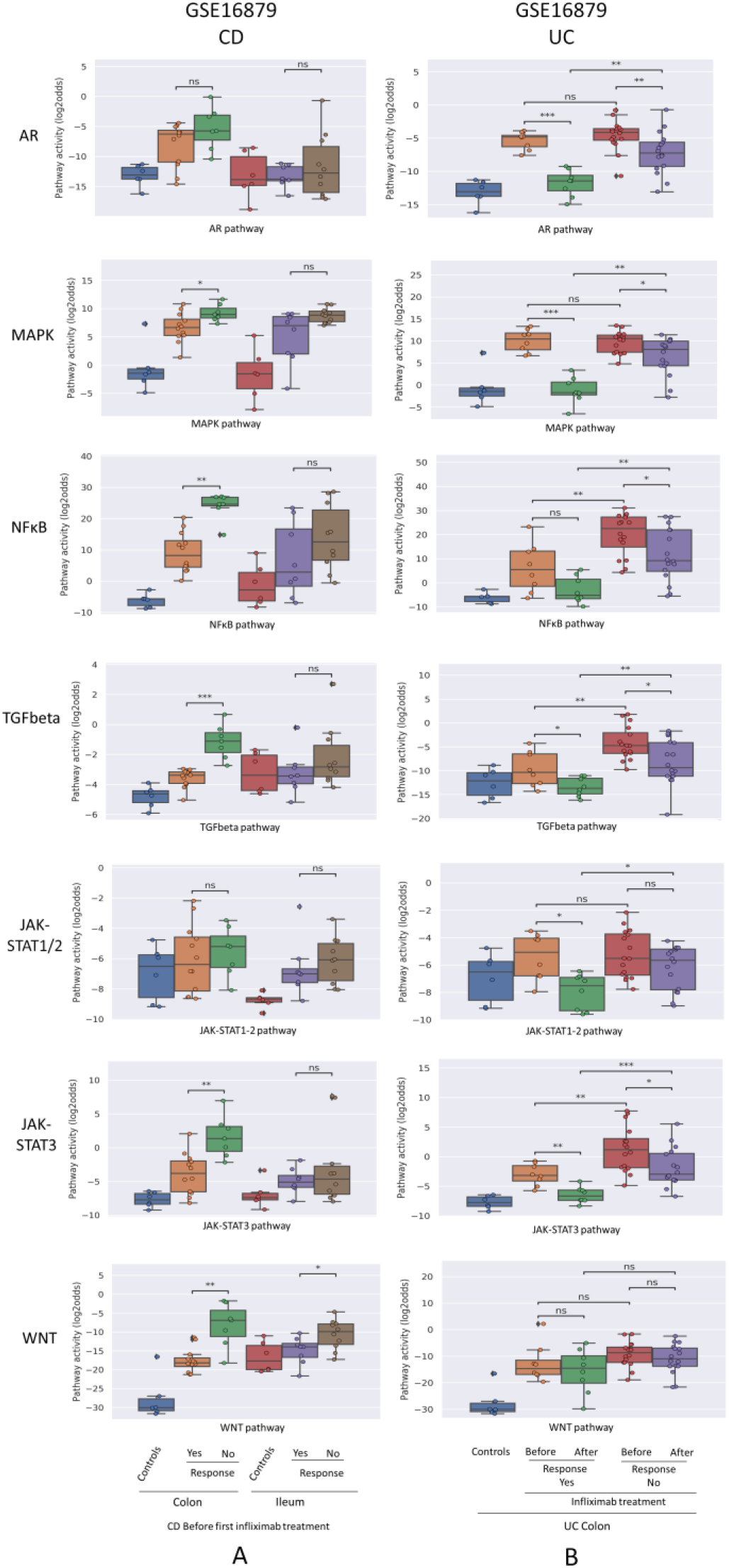
Prediction and assessment of response to anti-TNFα remission-induction treatment in CD (A) and UC (B). Datasets GSE16879. STP activity analysis of intestinal mucosa samples for prediction of response to anti-TNFα remission induction treatment in patients with CD and UC. STP PAS before and after remission induction treatment. For details, see Methods. STP PAS are shown for the AR, MAPK, NFκB, TGFβ, JAK-STAT1/2, JAK-STAT3, and Wnt STPs. Two sided Mann–Whitney–Wilcoxon statistical tests were performed; p-values are indicated in the figures as *p < 0.05, **p < 0.01, ***p < 0.001, ****p < 0.0001, ns: not significant. For supporting analysis results, see Supplementary Table 1.

In the second UC study (GSE23597, lesion-independent biopsy) two different dosages of anti-TNFα had been administered, and data were combined for our analysis. Again, high pre-treatment NFκB, TGFβ and JAK-STAT3 PAS were associated with resistance to remission-induction treatment (but not related to failure of maintenance treatment) (Figure 4; supplementary Table 1 and summary Table 1).

Summarizing these results, remission-induction response to the TNFα inhibitor Infliximab in patients with colon-localized CD and UC was reproducibly predicted by high pre-treatment NFκB, TGFβ, and JAK-STAT3 signa transduction pathway activity.

### Response monitoring of remission-induction and maintenance anti-TNFα treatment

Comparison of STP activity profiles between *post-treatment* responder and non-responder *groups* (not per individual patient) revealed the same hyperactive STPs which had been identified as pathogenic by comparing healthy individuals with IBD patients (Figure 2,3 and 4, Supplementary table 1 and summary Table 1, “*Patient group response assessment during treatment”*). Successful treatment resulted in return to (near) normal STP PAS values in the responder patient groups. Figure 4C illustrates the large differences in delta PAS induced by anti-TNFα treatment in responder patients compared to non-responder patients. These results support the pathogenic involvement of these STPs in IBD.

**Figure 3.**
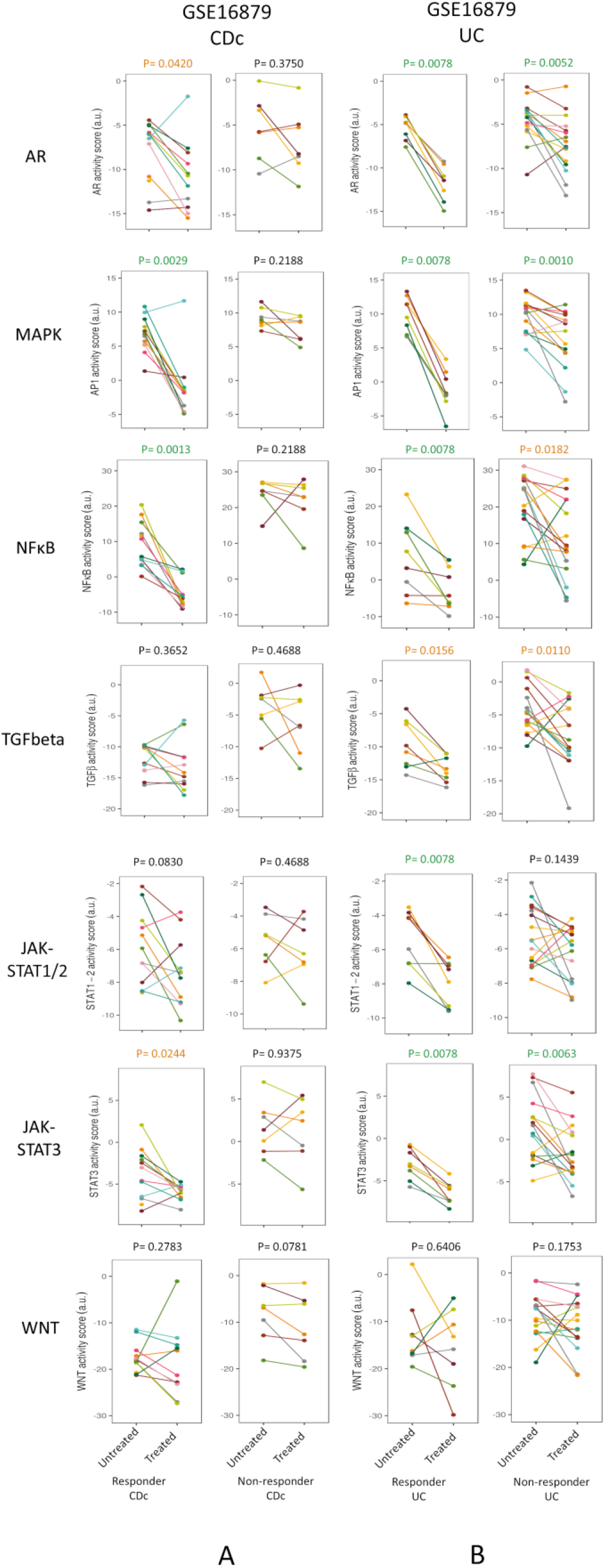
Prediction and assessment of response to anti-TNFα remission-induction treatment. Dataset GSE16879. STP activity analysis of intestinal mucosa samples for prediction of response to anti-TNFα remission induction treatment in patients with CDc (A) and UC (B). STP PAS before and after remission induction treatment, for responder and non-responder patients. For details, see Methods. STP PAS are shown for the AR, MAPK, NFκB, TGFβ, JAK-STAT1/2, JAK-STAT3, and Wnt STPs. From left to right: CDc responders; CDc non-responders; UC responders; UC non-responders. Pre-treatment (left) and post-treatment (right) PAS are connected by a line for each individual patient. Paired t-tests were performed; p-values are indicated above figures. For associated barplots, see Figure 2. For supporting analysis results, see Supplementary Table 1.

**Figure 4.**
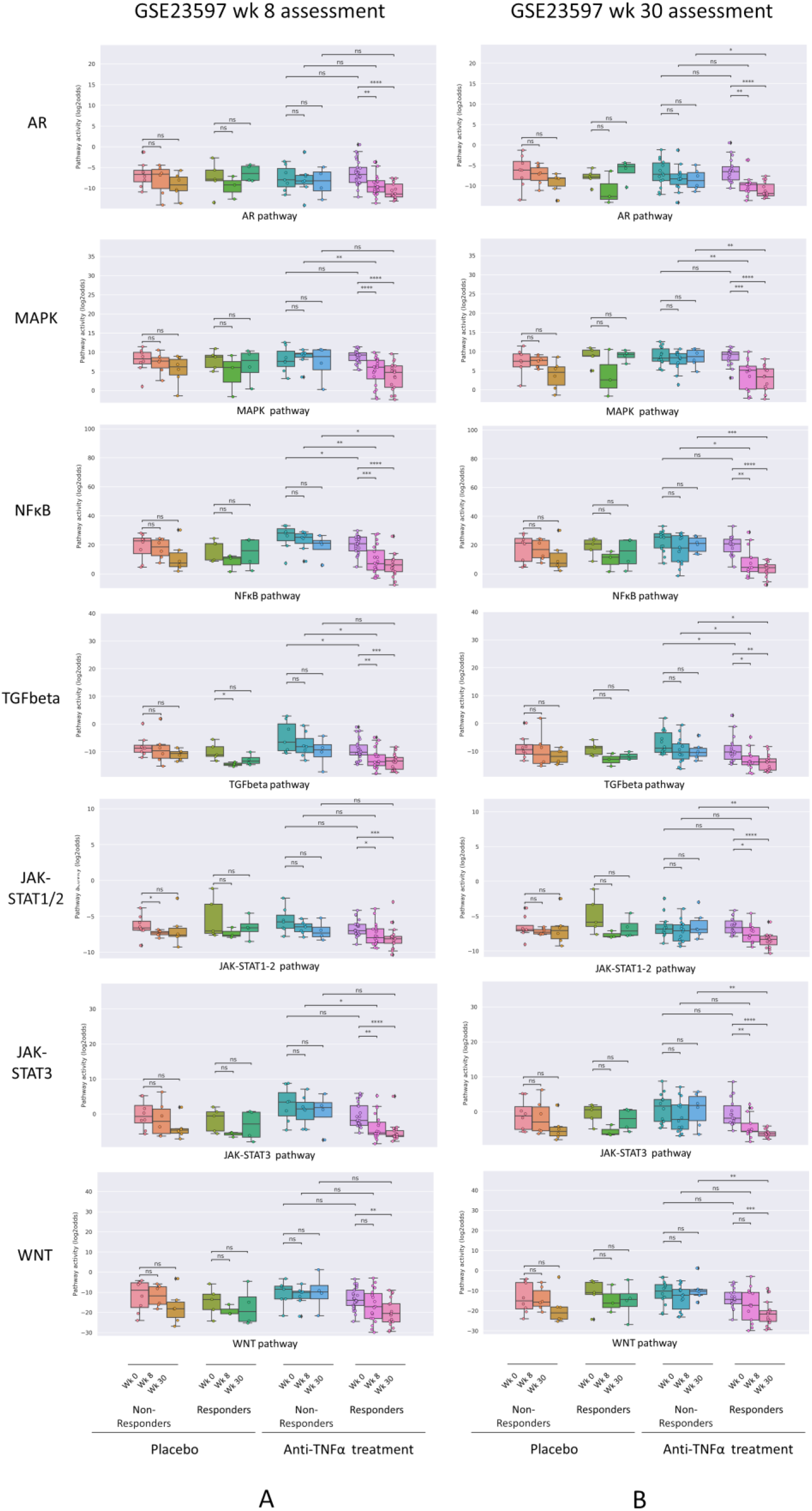
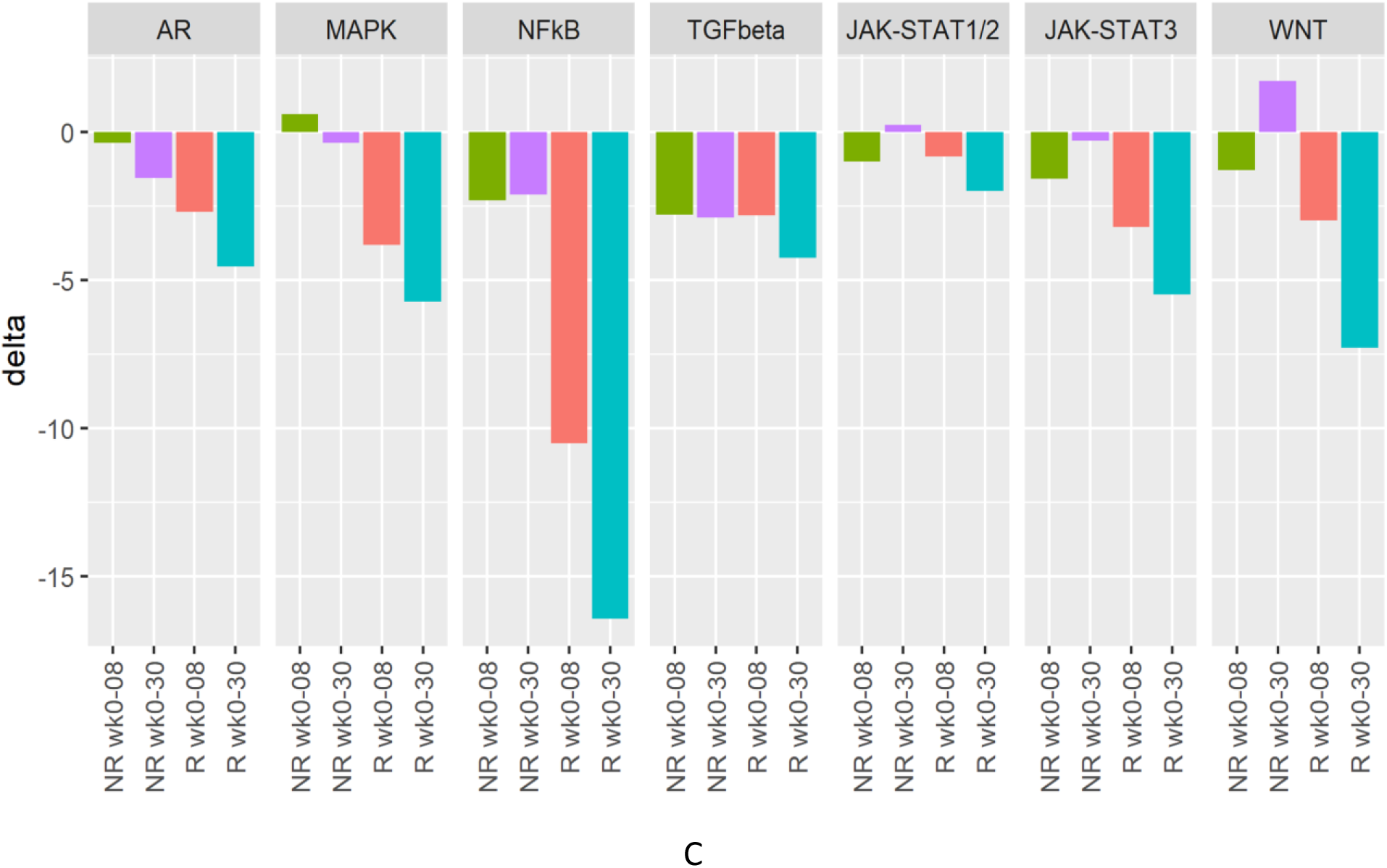
Assessment of response to anti-TNFα remission-induction versus maintenance treatment. Dataset GSE23597. STP activity analysis of intestinal mucosa samples for assessment of response to low or high dose anti-TNFα remission maintenance treatment of patients with UC. Clinical response to treatment was assessed at 8 (A, remission-induction treatment) and 30 (B, maintenance treatment) weeks after start of treatment. Treatment with infliximab, 5 or 10 mg/kg, or placebo. Colon mucosa samples: 0, 8, 30 weeks of treatment. For STP analysis, data of 5 and 10 mg/kg were combined. For details, see Methods. A: STP activity scores (PAS) at 0, 8, 30 weeks of treatment. Clinical response assessment at 8 weeks, following remission induction treatment. B. PAS at 0, 8, 30 weeks of treatment. Clinical response assessment at 30 weeks, following remission-induction and maintenance treatment. C. Mean delta (delta PAS calculated per individual patient) PAS between 0 and 8 weeks, and between 0 and 30 weeks of treatment, for responders (R; red and turquoise) and non-responders (NR; green and purple). STP PAS shown for AR, MAPK, NFκB, TGFβ, JAK-STAT1/2, JAK-STAT3, and Wnt STPs. PAS on Y-axis on a log2 odds scale. Two sided Mann–Whitney–Wilcoxon statistical tests were performed; p-values are indicated in the figures as *p < 0.05, **p < 0.01, ***p < 0.001, ****p < 0.0001, ns: not significant. For supporting analysis results, see Supplementary Table 1

Clinically it is expected to be of clinical relevance to know whether a patient has a biochemical response, for example to guide therapy decisions in case of an unclear clinical presentation. To check whether successful remission-induction can be assessed in an individual patient by comparing pre- and post-treatment STP activity profiles in sequentially taken mucosal biopsy samples, we calculated the delta STP PAS values for each individual patient (Figure 3, supplementary table 1; Table 1, *“Individual patient response assessment during treatment”*).

In individual UC patients who underwent successful remission-induction therapy (GSE16879), the decrease (delta) in AR and MAPK PAS was higher than in non-responder patients (supplementary Table 1). In addition, successful maintenance therapy in individual UC patients (GSE23597) was also characterized by a larger decrease (delta) in MAPK PAS (supplementary Table 1 and summary Table 1).

### Comparison of STP analysis with data analysis results obtained by the investigators who performed the clinical studies and generated the datasets

We compared our STP analysis results with the data analysis results obtained by the investigators who had generated the Affymetrix data which we analyzed for the purpose of the current study. For their data analysis they used conventional public data analysis tools and compared groups of samples, aiming to identify differentially involved cellular biochemical processes. In contrast to the current analysis, their analysis was not performed on individual samples. Below we briefly summarize their results with the used data analysis tools, per dataset.

GSE16879. Data was analyzed using Bioconductor tools, with focus on differentially involved antimicrobial peptides (AMP) genes. In non-responders to infliximab, differences in AMP gene expression were not found, neither before nor during treatment.

GSE52746. Differential expression and clustering analysis was performed using Ingenuity Pathway Analysis (IPA). Top 10 biochemical processes identified were: Docosahexanoic Acid signaling; IL17A signaling in fibroblasts; tumoricidal function of hepatic natural killer cells; role of IL17F in allergic inflammatory airway disease; role of IL17A in psoriasis; granulocyte adhesion and diapedesis; differential regulation of cytokine production in intestinal epithelial cells by IL17A and IL17F; agranulocyte adhesion and diapedesis; role of Tissue Factor in cancer.

Mechanistic networks: IL1B, IL17, NFkB, STAT3 are candidate upstream regulators. The investigators concluded that in non-responders an IL-17A-driven response was present which was not sensitive to anti-TNFα blockade. While involvement of NFκB and JAK-STAT3 pathways was identified, no information on activity status of these signaling pathways was obtained.

GSE23597. Data was analyzed using differential expression and clustering analysis with IPA. Top 6 biochemical processes identified were: cell-to-cell signaling and interaction; cellular movement; hematological system development and function; immune cell trafficking; inflammatory response; skeletal and muscular disorders. No information on involvement of specific signaling pathways and their activity was reported.

### Summary of results

Despite many differences between the investigated studies, STP analysis results were generally consistent across patient groups, both for CD and UC. We identified the MAPK STP as a ‘core’ abnormally active STP in IBD, while NFκB, TGFβ, JAK-STAT1/2, JAK-STAT3, and possibly the Wnt pathway, seem to be differentially involved in the various clinical IBD presentations. High activities of NFκB, TGFβ and JAK-STAT3 pathways prior to treatment were predictive for resistance to anti-TNFα treatment in patients with colon-localized IBD, but not in ileal CD. Successful remission-induction treatment and maintenance was associated with a reset in pathway activity towards normal. For all STPs there was a large variation in STP PAS among patients within the various IBD subtypes.

## Discussion

We previously reported a completely new method to quantitatively measure activity of clinically relevant signal transduction pathways (STP) on transcriptome data of individual cell-containing samples ^7^,^8^,^9^,^10^,^11^,^12^,^7^,^13^,^14^. Using this technology to analyze RNA expression data of colon mucosa biopsy samples, signaling pathway activity was investigated in CD and UC, and related to pathophysiology of IBD and the clinical response to anti-TNFα treatment.

Since abnormal activity of STPs is informative on the pathophysiology of a disease (the “mechanism of disease”) we investigated which STPs are abnormally active in different forms of IBD. Activity of the MAPK growth factor pathway was consistently increased in colon biopsies of patients with either UC or CD, irrespective of colonic or ileal disease location. Clinically effective anti-TNFα treatment (partly) corrected this abnormal signaling pathway activity, further supporting a core pathogenic role for the MAPK signaling pathway in IBD, independent of disease location. Indeed, the MAPK pathway has been described as an important signaling pathway in a number of related auto-immune diseases, such as IBD and rheumatoid arthritis ^22^,^23^,^24^,^25^.

A few signaling pathways showed differential activity between colon- and ileum-located IBD. Activity of the NFκB, AR, JAK-STAT3, Wnt, and (less consistently) the TGFβ, signaling pathways was increased in patients with IBD disease localization in the colon, while the JAK-STAT1/2 pathway was specifically active in ileal localized CD, suggesting differences in pathophysiology depending on disease location. This is in line with current thinking regarding differences between ileal- and colon-localized IBD, including marked differences in clinical response to anti-TNFα treatment ^18^. The inflammatory NFκB pathway, driving production of TNFα, is well known as a core mechanistic pathway in colon-but less in ileum-located IBD ^26^. Indeed, patients with ileum-located CD generally are known to respond less well to anti-TNFα treatment ^27^. Due to sequential pathway activation (e.g. an active NFκB pathway produces IL6, resulting in activation of the JAK-STAT3 pathway) and intracellular crosstalk between signaling pathways (e.g. cooperation between transcription factors of MAPK and TGFβ pathways), multiple pathways, i.e. the Wnt, MAPK, JAK-STAT and TGFβ pathways, are recruited in the pathogenic process underlying IBD ^28^,^29^,^30^,^31^,^32^,^33^. While most of these pathways are thought to contribute to the inflammatory character of the disease, the TGFβ pathway may play an instrumental role in the fibrotic process associated with IBD ^34^.

Our results support the increasing evidence that the pathogenesis (or *mechanism of disease*) of ileal IBD is distinct from colon-located IBD ^35^,^36^,^37^,^38^,^39^,^40^,^41^. Interferons are activators of the JAK-STAT1/2 pathway which was in our analysis identified as specifically active in ileal CD ^42^. Indeed, interferon lambda concentrations have been reported as increased in mucosa of IBD-affected ileum, both in mouse models and in patients, supporting our findings ^43^. We believe that STP analysis contributes to our understanding of the *mechanism of disease* underlying the various manifestations of IBD and may improve diagnosis and differential diagnosis between colon- and ileum-located disease.

In patients with a diagnosis of IBD, either CD or UC, a biomarker assay to predict response to anti-TNFα treatment is needed because of potentially serious side effects of the treatment and significant treatment costs ^6^. In our study, in IBD patients with colon involvement, high activity of NFκB, JAK-STAT3 and TGFβ pathways prior to treatment was associated with failure to induce remission by anti-TNF-α treatment. The NFκB pathway is the prime target for anti-TNFα treatment, since activity of this pathway is both induced by TNFα and responsible for production of TNFα ^27^,^26^. The known secondary activation of JAK-STAT3 and TGFβ pathways by a (TNFα-)activated NFκB pathway readily explains their combined association with resistance to anti-TNFα treatment ^28^,^29^,^30^.

NFκB pathway activity seemed to play a less dominant role in ileal-CD, clearly in line with anti-TNFα treatment being less effective in patients with ileal-restricted CD ^27^. Indeed, in the here analyzed clinical study, clinical response to anti-TNFα treatment had been separately defined on less strict criteria than used for clinical response in the other IBD groups ^18^. Together our results provide initial evidence that STP analysis can be used to predict response to anti-TNFα treatment, except in patients with ileum-restricted CD.

While considered highly necessary from a health economic standpoint as well as from a clinical point of view, currently clinical response to anti-TNFα treatment cannot be predicted in IBD patients ^44,45^,^46^,^6, 47^. Literature data report that 20–40% of patients in clinical trials and 10–20% of patients in “real life” series are primary resistant to anti-TNFα induction therapy, while secondary resistance after a year treatment is even more common ^48^.

Our results suggest that to overcome resistance to anti-TNFα treatment, it may be needed to target the NFκB signal transduction pathway in a different manner than by inhibiting its ligand TNFα ^49^,^50^. Indeed, the signaling pathways here identified as abnormally active in IBD patients provide new drug targets and opportunities for alternative treatment strategies. Development of MAPK pathway inhibitors for IBD has been explored without clinical success, probably due to the complex and not well understood role of this pathway in IBD ^23^,^22^,^25, 24^. JAK-STAT pathway inhibitors are being explored for IBD and the JAK-STAT3 pathway inhibitor tofacitinib has been approved for treatment of UC ^31^,^51^,^7^,^52^,^53^,^54^. TGFβ inhibitors are predominantly being developed for cancer immunotherapy, but reducing intestinal fibrosis in IBD has been suggested as a potential clinical application ^53^. For ileum-localized CD, based on our current results we hypothesize that JAK-STAT1/2 inhibitors may be more effective than TNFα inhibitors ^55^,^31^.

Potentially relevant for any treatment of IBD is the observation that activity of the signaling pathways showed large variations among patients with IBD. This suggests that clinical efficacy of a drug, including an anti-TNFα drug, in an individual patient may depend on the level of activity of the targeted signaling pathway, implicating that a personalized treatment approach is likely to be necessary to optimize treatment results. This requires an assay to measure STP activity in a sample of every individual patient to predict response to a drug. For example, a decision on treatment with an anti-TNFα drug such as infliximab ideally would be preceded by measuring NFκB pathway activity to guide this treatment decision.

For each Affymetrix dataset for which we performed STP analysis, data-analysis results obtained by the original investigators were compared to our results. While gene expression profiles had been obtained for CD and UC patient groups (instead of individual patients), no gene profile had been identified by the previous investigators to separate anti-TNFα responder and non-responder group, and no information on STP activity had been extracted from the Affymetrix data. This emphasizes the difference between our STP activity analysis, which can be performed on individual patient samples and provides clinically actionable STP information for an *individual patient* (e.g. responder or non-responder to infliximab), and other bioinformatics tools which are used to discover gene expression profiles as potential novel biomarkers for *groups of patients*.

In conclusion, STP analysis provided a first-of-a-kind insight into the mechanism of disease of IBD and its relation to treatment resistance. Our results provide new treatment targets, for example the MAPK pathway in all types of IBD, in ileal Crohn’s disease the JAK-STAT1/2 pathway, and in patients refractory to anti-TNFα treatment the NFκB, TGFβ, and JAK-STAT3 pathways.

## Data Availability Statement

All datasets presented in this study are publicly available from the Gene Expression Omnibus (GEO) database ^15^.

## Author Contributions

WB: Data, results analysis, concept and writing. WV: pathway model development, results analysis. AS: pathway model development, concept, results analysis, and writing. All authors contributed to the article and approved the submitted version.

## Acknowledgments

We wish to acknowledge all investigators who performed the clinical studies and generated the publicly available GEO datasets used in the current analysis.

