## Supplemental figures for "Improved diagnosis of inflammatory bowel disease and prediction and monitoring of response to anti-TNF alpha treatment based on measurement of signal transduction pathway activity"

Philips Research, Eindhoven, The Netherlands

**Conflict of interest statement: All authors are regular employees of Philips**

### Supplemental figures:

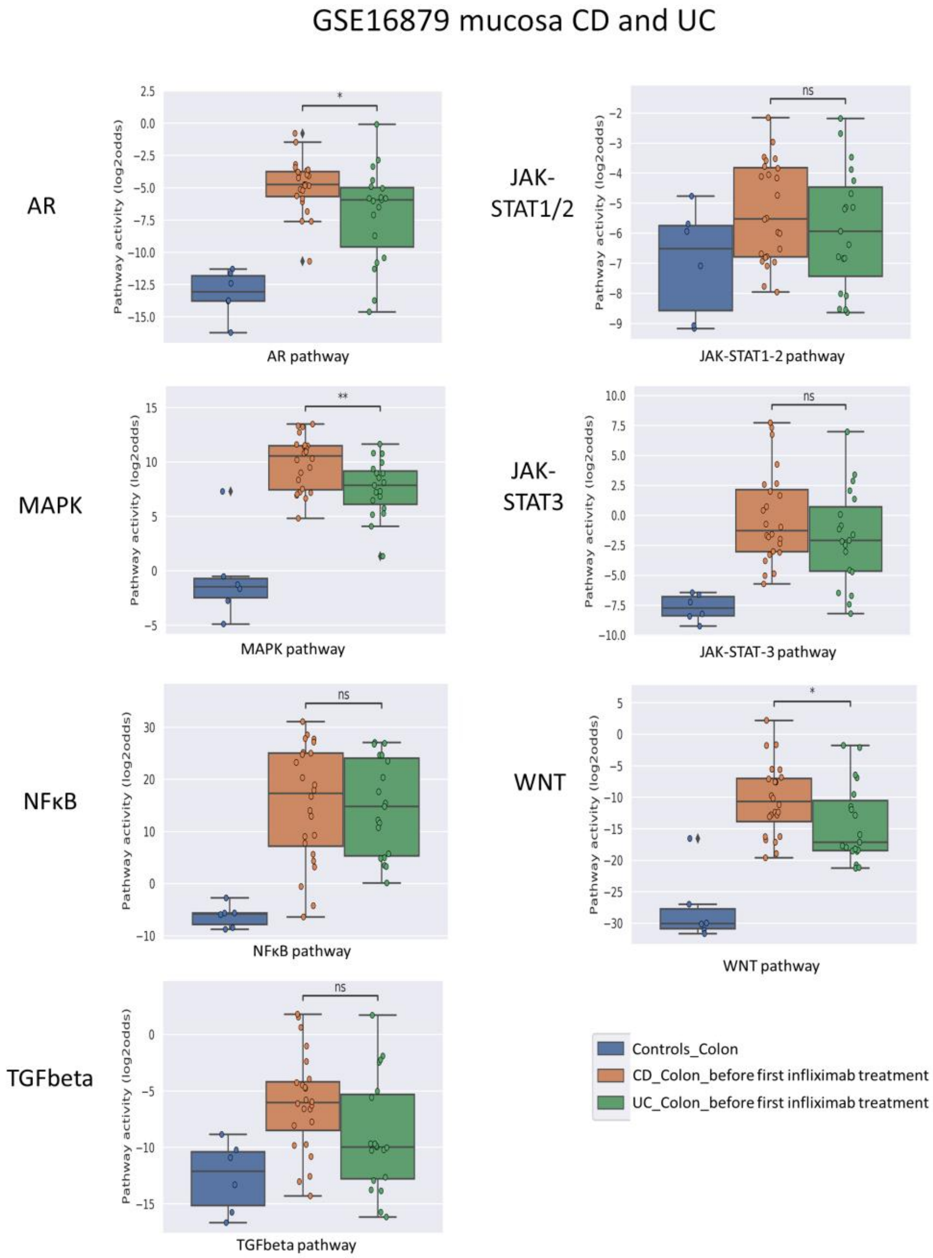


Figure S1. **Differential diagnosis of CD and UC.** Dataset GSE16879. Illustration of differences in STP PAS between CD and UC, irrespective of response to anti-TNFα treatment. Mucosa samples are from healthy control individuals and pre-treatment mucosa samples from IBD patients. For details, see Methods. Signal transduction pathway (STP) activity scores (PAS) are shown (A-G) for the androgen receptor (AR), MAPK , NFκB, TGFβ, JAK-STAT1/2 (STAT1-2), JAK-STAT3 (STAT3), and Wnt STPs. PAS on Y-axis on a log2 odds scale. From left to right: Healthy control, CD, UC. Two sided Mann-Whitney-Wilcoxon statistical tests were performed; p-values are indicated in the figures as $*p < 0.05$, $**p < 0.01$, ns: not significant.

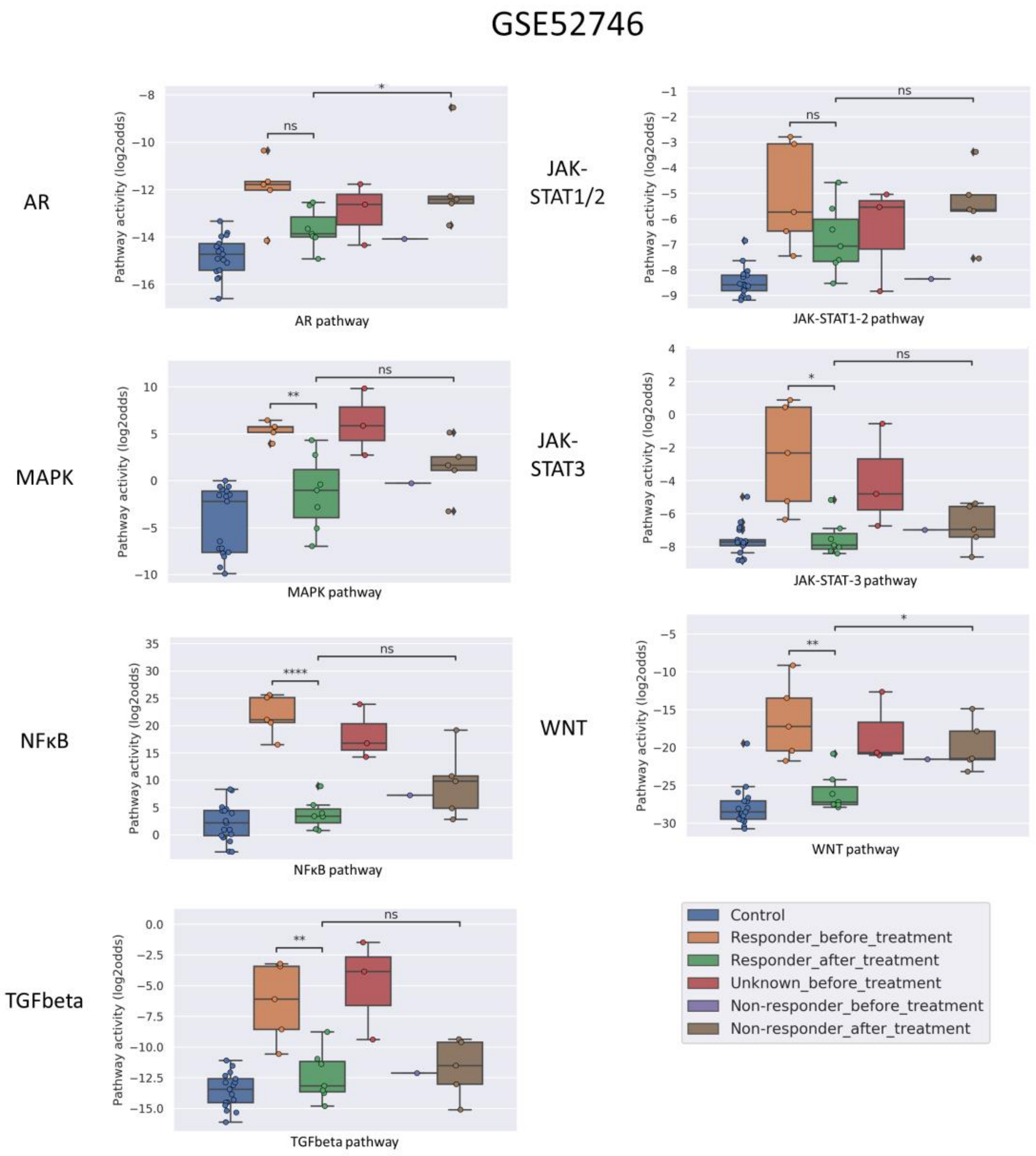


Figure S2: **Prediction and assessment of response to anti-TNFα remission-induction treatment in CD.** Dataset GSE52746 on top. STP activity analysis of intestinal mucosa samples for prediction of response to anti-TNFα remission induction treatment in patients with CD. STP PAS before and after remission induction treatment, for responder and non-responder patients. For some patients only a sample before treatment was taken. For details, see Methods. STP PAS are shown for the androgen receptor (AR), MAPK , NFκB, TGFβ, JAK-STAT1/2 (STAT1-2), JAK-STAT3 (STAT3), and Wnt STPs. PAS on Y-axis on a log2 odds scale. From left to right: Dataset GSE52746: Healthy control, pretreatment samples, response unknown; responders before and after treatment; non-responders before and after treatment; dataset Two sided T-test statistical tests were performed; p-values are indicated in the figures as *p < 0.05, **p < 0.01, ****p < 0.0001, ns: not significant.
